# Parental awareness and attitudes towards prevention of respiratory syncytial virus in infants and young children in Australia

**DOI:** 10.1101/2023.10.18.23297187

**Authors:** Charlie Holland, Megan Baker, Amber Bates, Catherine Hughes, Peter C Richmond, Samantha Carlson, Hannah C Moore

## Abstract

**Aims:** To assess parental awareness of respiratory syncytial virus (RSV) and the level of acceptance of future RSV prevention strategies.

**Methods:** A cross-sectional online survey was implemented targeting “future” and “current” parents of children aged ≤5 years in Australia.

**Results:** From 1,992 eligible participants, two non-mutually exclusive subgroups were formed; “current” parents (N=1931) and “pregnant/planning” parents (N=464; 403 also “current” parents; 61 “future” parents). Participants were predominantly (86.6%) aged 25-39 years and 68.5% with university education. The majority (89.6% current; 78.7% future) had heard of RSV. Of those, 64.2% (current) and 50.0% (future) were aware that pneumonia is associated with RSV; 71.8% (current) and 52.1% (future) were aware that bronchiolitis is associated. In multivariable logistic regression analyses, Australian-born parents (aOR=2.47 [95%CI:1.48-4.12]), living in the Eastern States (e.g., New South Wales: aOR=6.15 [95%CI:2.10-18.04]), with a university level education (aOR=2.61 [95%CI:1.38-4.94]) and being a current parent (aOR=12.26 [95%CI:2.82-53.28]) were associated with higher RSV awareness. There was a high level of acceptance for maternal vaccines (future: 79.3%) and infant immunisation (all: 81.7%).

**Conclusion:** While RSV awareness and immunisation acceptance was high, there was limited knowledge of severity of RSV, especially in future parents. Education campaigns need to be developed to increase RSV knowledge.

**Key notes:** The success of RSV immunisation programs is dependent on the community having sufficient disease awareness and acceptance of new immunisation strategies. Majority of parents in the study had heard of RSV, however lacked awareness on associated conditions and disease severity, particularly future parents. Education and awareness campaigns are required to inform parents about RSV and future immunisations; there is a critical opportunity to increase awareness prior to immunisation arrival.

## Introduction

Respiratory syncytial virus (RSV) is the predominant cause of respiratory infections and is most prevalent in infants and young children.^1^ RSV causes 33.0 million annual cases globally, 3.6 million hospitalisations and 101,400 deaths in children aged less than 5 years.^2^ RSV is the leading cause of bronchiolitis and pneumonia, and is associated with later respiratory morbidity including asthma and long-term wheezing.^3,4,5^ Infants at risk of developing severe RSV infections include those <6 months or born prematurely (<37 weeks gestation), and children of all ages with comorbidities.^6,7^ RSV causes seasonal epidemics, often peaking in the winter months,^8^ but there has been some variability to the typical seasonal pattern since COVID-19.^9,10^ Currently, Palivizumab is the only licensed RSV prevention product,^11^ available to high-risk infants and requires costly monthly injections.^12,13^

Since 2019, the World Health Organization (WHO) has recognised RSV as a priority target for prevention through vaccination.^14^ The RSV vaccination landscape is rapidly progressing, with >30 maternal vaccine and single dose long-acting monoclonal antibody (mAb) candidates in clinical trials. Whilst no vaccine candidates have yet been approved for use in Australia, a single dose long-acting mAb has been approved in Europe and the United Kingdom.^15,16^ Thus, with the imminent arrival of RSV immunisation strategies in Australia, it is critical to understand what factors may contribute to uptake of these products. A Melbourne based study in 2019 found 83.0% of pregnant mothers had never heard of RSV before, though after receiving information, 77.0% were very likely to accept an RSV maternal vaccine.^17^ Similar findings were found in a recent global study.^18^ These findings suggest that maternal vaccine and mAb uptake may rely on parents having sufficient awareness and knowledge of RSV.^17,18^ There is currently a lack of contemporary data on RSV community awareness in Australia. We aimed to assess parent awareness of RSV and the level of acceptance of RSV prevention strategies.

## Methods

### Study design

We conducted an Australian wide cross-sectional survey from 15^th^ July to 27^th^ August 2022. Eligible participants were those aged ≥18 years, residing in Australia, could read and understand English and were either currently pregnant, planning to become pregnant in the next 6 months, a partner of a pregnant person, and/or were caring for at least one child aged ≤5 years. Eligible participants were divided into two non-mutually exclusive subgroups; 1) pregnant/planning, and 2) current parent. Participants who only fit into the pregnancy/planning subgroup formed an additional subgroup “future” parents which was used for analysis to compare future and current parents.

### Survey Instrument

Survey items were derived from previously published studies on childhood and pregnancy vaccine attitudes,^17,19^ as well as the research team’s expertise on RSV disease and immunisation strategies. Items were reviewed by our consumer representative for relevance to our intended population. The survey was refined through user testing by the research team. The final survey included 47 items focusing on awareness of childhood respiratory illnesses, awareness of RSV and willingness to accept and potentially pay for RSV immunisations strategies. The pregnant/planning subgroup were questioned on their acceptance towards RSV maternal immunisations and what source they refer to for information regarding infectious diseases and vaccines in pregnancy. The current parent subgroup were questioned on their acceptance towards RSV childhood immunisation and what information sources they refer to for infectious diseases and childhood vaccines. Participants fitting into both subgroups were questioned on both immunisation strategies and information sources. Following these questions, participants were provided with information on RSV and then asked the same questions on immunisation acceptance to determine if there were any changes to their responses. Participants were asked an open-ended question regarding what information they would need to decide on RSV prevention products.

### Recruitment

Online recruitment occurred through two avenues: promotion through community networks (Immunisation Foundation of Australia, a community-member network that advocates for immunisations; Tiny Sparks WA, a not-for-profit organisation for high-risk pregnancies and sick or premature infants) and targeted advertising on social media (Facebook, Instagram) facilitated by the digital team at Telethon Kids Institute. Following eligibility screening and consent, participants were directed to the web-based survey on REDCap.

### Statistical analysis

Descriptive statistics including frequencies with 95% confidence intervals (CI) were used for categorial responses, with comparison of proportions using chi-squared tests. To assess the predictors of RSV awareness, we used multivariable logistic regression with RSV awareness as a binary outcome (yes/no) with demographic and questionnaire responses as possible covariates. We present odds ratios (ORs) with 95% CI. Factors with a p-value of <0.2 in univariate analysis were included in a multivariable model. To assess for selection bias in the survey sample, the socio-demographic characteristics of respondents were compared to Australia’s general population using Australian Bureau of Statistics (ABS) data. STATA version 17 was used for all analyses. Open-ended question data were thematically analysed using the Braun and Clarke method^20^ to describe themes. Two researchers (CH and MB) independently applied initial codes following data familiarisation. Initial codes were then compared and manually collated into themes until consensus between both researchers was reached. Ethics approval was obtained by the University of Western Australia Human Research Ethics Committee (ET000182).

## Results

From a 6-week period, 1,992 eligible participants were recruited; 464 formed the pregnant/planning subgroup (61 participants who fit only into this subgroup were considered “future parents”) and 1,931 formed the current parent subgroup (403 participants fit into both subgroups). The majority of participants were aged 25-39 years (86.6%), were female (98.9%), born in Australia (77.2%) and spoke English as their primary language (85.1%; Table 1). Participants were distributed from all states of Australia, with New South Wales (27.9%), Western Australia (24.7%) and Victoria (22.8%) having the highest number of respondents. Majority of participants had a university degree (68.5%) and nearly half (49.3%) were working either full time or part-time. This was a high proportion compared to the ABS 2021 Census that reported 30.7% of the population have a university degree and 43.5% work full time or part-time.^21^

**Table 1.** Demographics of participants.

| Characteristic | Category | Future Parent<br>N=61 |  | Current Parent<br>N=1931 |  |
| --- | --- | --- | --- | --- | --- |
|  |  | n | (%) | n | (%) |
| **Age (years)** | 18-24 | 4 | 6.6% | 37 | 1.9% |
|  | 25-39 | 52 | 85.2% | 1673 | 86.6% |
|  | 40-59 | 5 | 8.2% | 221 | 11.4% |
| **Gender** | Female | 59 | 96.7% | 1911 | 99.0% |
|  | Male | 1 | 1.6% | 17 | 0.9% |
|  | Non-binary | 1 | 1.6% | 3 | 0.2% |
| **State** | Northern Territory | 1 | 1.6% | 7 | 0.4% |
|  | New South Wales | 20 | 32.8% | 535 | 27.7% |
|  | Australian Capital Territory | 1 | 1.6% | 41 | 2.1% |
|  | Victoria | 12 | 19.7% | 443 | 22.9% |
|  | Queensland | 8 | 13.1% | 280 | 14.5% |
|  | South Australia | 1 | 1.6% | 111 | 5.75% |
|  | Western Australia | 16 | 26.2% | 477 | 24.7% |
|  | Tasmania | 2 | 3.3% | 37 | 1.9% |
| **Country of birth** | Australia | 45 | 73.8% | 1492 | 77.3% |
|  | Other than Australia | 9 | 14.8% | 257 | 13.3% |
|  | No response | 7 | 11.5% | 182 | 9.4% |
| **Indigenous Status** | Non-Indigenous | 54 | 88.5% | 1720 | 89.1% |
|  | Indigenous | 0 | 0.0% | 25 | 1.3% |
|  | Prefer not to answer | 0 | 0.0% | 3 | 0.2% |
|  | No response | 7 | 11.5% | 183 | 9.5% |
| **Primary Language** | English | 51 | 83.6% | 1644 | 85.1% |
|  | Other than English | 3 | 4.9% | 100 | 5.2% |
|  | No response | 7 | 11.5% | 187 | 9.7% |
| **Highest Level of Education** | University | 46 | 75.4% | 1318 | 68.3% |
|  | TAFE/Apprenticeship | 5 | 8.2% | 257 | 13.3% |
|  | High school | 3 | 4.9% | 151 | 7.8% |
|  | Prefer not to answer | 0 | 0.0% | 12 | 0.6% |
|  | Other | 0 | 0.0% | 10 | 0.5% |
|  | No response | 7 | 11.5% | 183 | 9.5% |
| **Employment Status** | Full time/Part time | 39 | 63.9% | 943 | 48.9% |
|  | Maternity leave | 10 | 16.4% | 550 | 28.5% |
|  | Home duties | 1 | 1.6% | 216 | 11.2% |
|  | Unemployed | 0 | 0.0% | 9 | 0.5% |
|  | Student | 2 | 3.3% | 28 | 1.5% |
|  | Prefer not to answer | 2 | 3.3% | 3 | 0.2% |
|  | No response | 7 | 11.5% | 182 | 9.4% |

Most participants (>80.0%) in the future and current parent subgroups had heard of all the childhood respiratory illnesses and pathogens, aside from *Bordetella pertussis* where only half of participants indicated they were aware (50.8% future: 56.3% current; Table 2). This was despite the majority of participants having heard of whooping cough (95.1% future: 95.0% current). Over three-quarters (78.7%) of future and 89.6% of current parents reported awareness of RSV. Notably, awareness was lower in the future parent subgroup compared to the current parent subgroup for all conditions; but only reached statistical significance for RSV, bronchiolitis and pneumonia (Table 2).

**Table 2.** Participant awareness of respiratory illnesses and pathogens (N=1992).

| Childhood Respiratory Condition | Response | Future Parent Subgroup<br>N=61 |  | Current Parent Subgroup<br>N=1931 |  | p-value |
| --- | --- | --- | --- | --- | --- | --- |
|  |  | n | (%) | n | (%) |  |
| Childhood Respiratory Pathogen: |  |  |  |  |  |  |
| Influenza (the Flu) | Yes | 58 | 95.1% | 1835 | 95.0% | 0.94 |
|  | No | 0 | 0.0% | 4 | 0.2% |  |
|  | No response | 3 | 4.9% | 92 | 4.8% |  |
| Whooping cough | Yes | 58 | 95.1% | 1835 | 95.0% | 0.94 |
|  | No | 0 | 0.0% | 4 | 0.2% |  |
|  | No response | 3 | 4.9% | 93 | 4.8% |  |
| Chest infections | Yes | 56 | 91.8% | 1832 | 94.9% | 0.18 |
|  | No | 1 | 1.6% | 6 | 0.3% |  |
|  | No response | 4 | 6.6% | 93 | 4.8% |  |
| Pneumonia | Yes | 56 | 91.8% | 1826 | 94.6% | 0.008 |
|  | No | 2 | 3.3% | 8 | 0.4% |  |
|  | No response | 3 | 4.9% | 97 | 5.0% |  |
| Wheezing | Yes | 52 | 85.2% | 1770 | 91.7% | 0.094 |
|  | No | 5 | 8.2% | 63 | 3.3% |  |
|  | No response | 4 | 6.6% | 98 | 5.1% |  |
| Bronchiolitis | Yes | 51 | 83.6% | 1767 | 91.5% | 0.025 |
|  | No | 6 | 9.8% | 66 | 3.4% |  |
|  | No response | 4 | 6.6% | 98 | 5.1% |  |
| COVID-19 | Yes | 57 | 93.4% | 1834 | 95.0% | 0.80 |
|  | No | 0 | 0.0% | 3 | 0.2% |  |
|  | No response | 4 | 6.6% | 94 | 4.9% |  |
| Common cold | Yes | 58 | 95.1% | 1833 | 94.9% | 0.96 |
|  | No | 0 | 0.0% | 0 | 0.0% |  |
|  | No response | 3 | 4.9% | 98 | 5.1% |  |
| Asthma | Yes | 57 | 93.4% | 1829 | 94.7% | 0.84 |
|  | No | 0 | 0.0% | 3 | 0.2% |  |
|  | No response | 4 | 6.6% | 99 | 5.1% |  |
| Ear infections | Yes | 58 | 95.1% | 1826 | 94.6% | 0.86 |
|  | No | 0 | 0.0% | 0 | 0.0% |  |
|  | No response | 3 | 4.9% | 105 | 5.4% |  |
| Childhood Respiratory Pathogen: |  |  |  |  |  |  |
| Influenza virus | Yes | 57 | 93.4% | 1830 | 94.8% | 0.30 |
|  | No | 1 | 1.6% | 7 | 0.4% |  |
|  | No response | 3 | 4.9% | 94 | 4.9% |  |
| Bordetella pertussis | Yes | 31 | 50.8% | 1087 | 56.3% | 0.48 |
|  | No | 27 | 44.3% | 714 | 37.0% |  |
|  | No response | 3 | 4.9% | 130 | 6.7% |  |
| Respiratory Syncytial Virus (RSV) | Yes | 48 | 78.7% | 1731 | 89.6% | <0.001 |
|  | No | 10 | 16.4% | 97 | 5.0% |  |
|  | No response | 3 | 4.9% | 103 | 5.3% |  |
| **SARS-CoV-2** | Yes | 55 | 90.2% | 1757 | 91.0% | 0.94 |
|  | No | 3 | 4.9% | 78 | 4.0% |  |
|  | No response | 3 | 4.9% | 96 | 5.0% |  |
| **Rhinovirus** | Yes | 51 | 83.6% | 1694 | 87.7% | 0.32 |
|  | No | 7 | 11.5% | 127 | 6.6% |  |
|  | No response | 7 | 11.5% | 127 | 6.6% |  |

\*\*Subgroups used for analysis were the future parent subgroup (participants who do not currently have any young children) and the current parent subgroup (participants who do currently have young children).

Pneumonia and whooping cough caused the highest level of concern (Figure 1). Respondents with RSV awareness had limited knowledge of associated RSV conditions (Supplementary Table 1). Only half (50.0%) of future parents and 64.2% of current parents were aware that pneumonia is associated with RSV; and 52.1% of future parents and 71.8% of current parents were aware that bronchiolitis is associated. Parents also demonstrated a low level of understanding that asthma (31.3% future; 30.6% current) and wheezing (56.3% future; 74.7% current) can be associated with RSV. From 1,931 current parents, 712 (36.9%) indicated that at least one of their children aged ≤5 years had been infected with RSV before; of those, 304 (42.3%) reported their child required hospitalisation.

**Figure 1.**
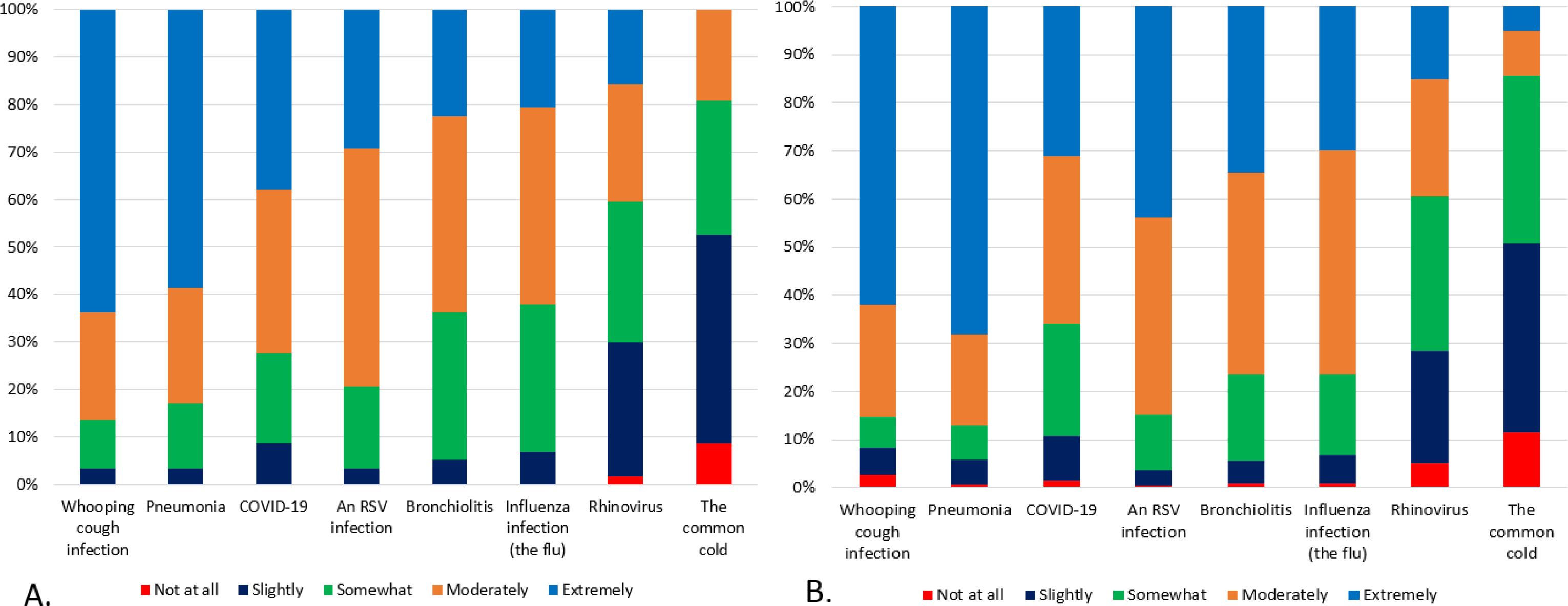
Level of worry towards childhood respiratory illnesses in the future parent subgroup (A) and current parent subgroup (B). Note: excludes participants who did not respond. Participants were asked about how worried they would be if their young child were to contract such conditions. Conditions are listed in order of level of worry (from left to right).

Univariate logistic regression analysis suggested being born in Australia, speaking English as a primary language, living in the eastern states, having a university degree, and working full time or part time were associated with a higher level of awareness of RSV (Table 3). In the adjusted model, demographic factors associated with RSV awareness were being born in Australia (aOR=2.47; 95%CI: 1.48-4.12), English as a primary language (aOR=1.37; 95%CI: 0.62-3.05) and living in the Eastern states. Compared to future parents, current parents had a 12-fold increased likelihood of RSV awareness (aOR=12.26; 95%CI: 2.82-52.28). Parents with personal experience of RSV were 6.6 times more likely to be aware of RSV (aOR=6.66, 95%CI: 3.26-13.64).

**Table 3.**
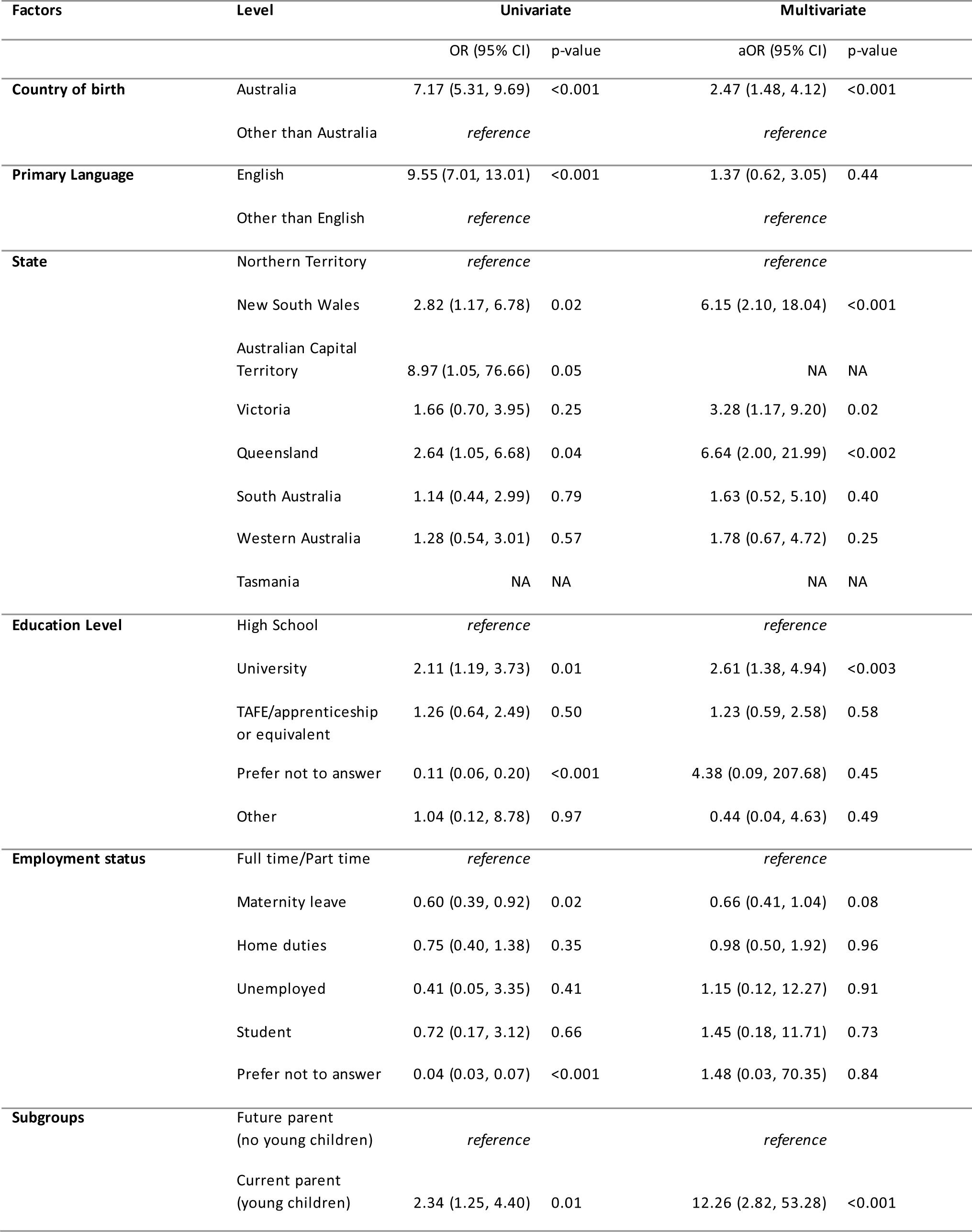
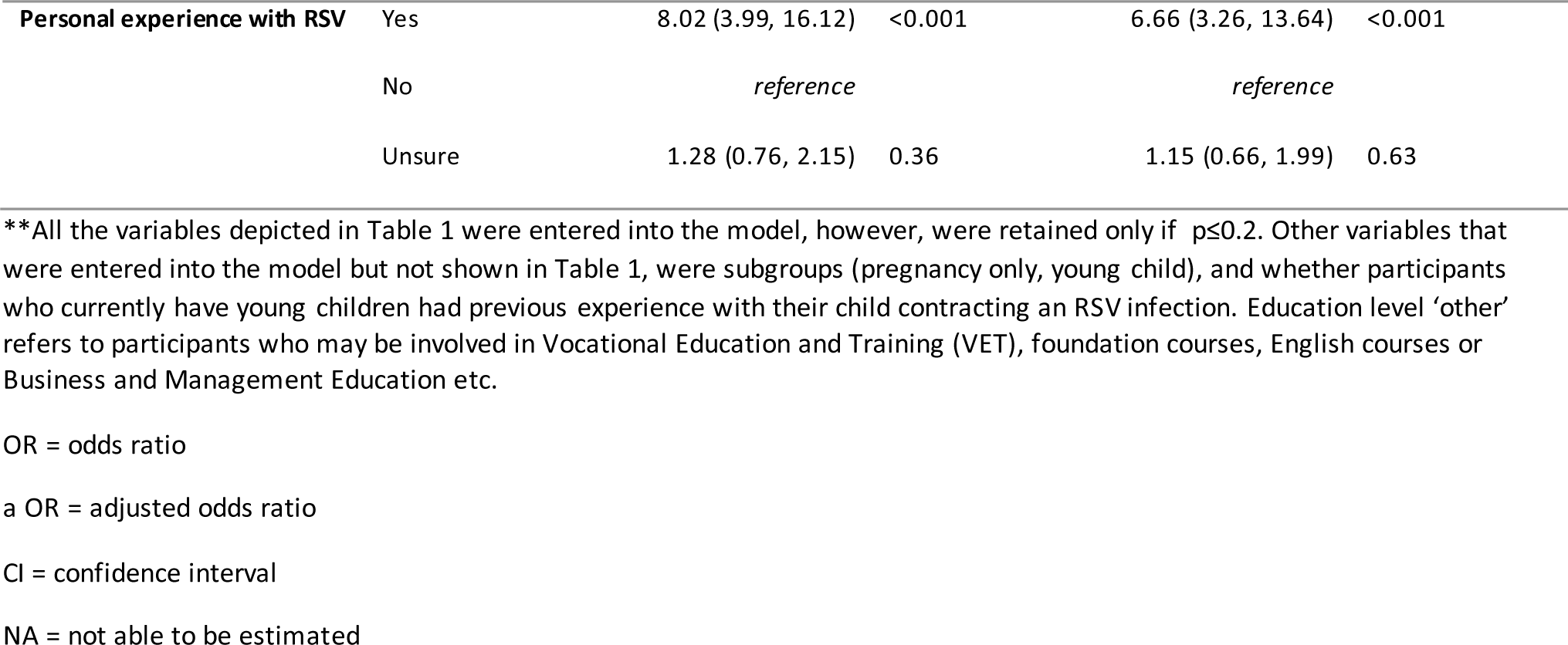
Factors associated with awareness of RSV.

| Factors | Level | Univariate |  | Multivariate |  |
| --- | --- | --- | --- | --- | --- |
|  |  | OR (95% CI) | p-value | aOR (95% CI) | p-value |
| Country of birth | Australia | 7.17 (5.31, 9.69) | <0.001 | 2.47 (1.48, 4.12) | <0.001 |
|  | Other than Australia | reference |  | reference |  |
| Primary Language | English | 9.55 (7.01, 13.01) | <0.001 | 1.37 (0.62, 3.05) | 0.44 |
|  | Other than English | reference |  | reference |  |
| State | Northern Territory | reference |  | reference |  |
|  | New South Wales | 2.82 (1.17, 6.78) | 0.02 | 6.15 (2.10, 18.04) | <0.001 |
|  | Australian Capital Territory | 8.97 (1.05, 76.66) | 0.05 | NA | NA |
|  | Victoria | 1.66 (0.70, 3.95) | 0.25 | 3.28 (1.17, 9.20) | 0.02 |
|  | Queensland | 2.64 (1.05, 6.68) | 0.04 | 6.64 (2.00, 21.99) | <0.002 |
|  | South Australia | 1.14 (0.44, 2.99) | 0.79 | 1.63 (0.52, 5.10) | 0.40 |
|  | Western Australia | 1.28 (0.54, 3.01) | 0.57 | 1.78 (0.67, 4.72) | 0.25 |
|  | Tasmania | NA | NA | NA | NA |
| Education Level | High School | reference |  | reference |  |
|  | University | 2.11 (1.19, 3.73) | 0.01 | 2.61 (1.38, 4.94) | <0.003 |
|  | TAFE/apprenticeship or equivalent | 1.26 (0.64, 2.49) | 0.50 | 1.23 (0.59, 2.58) | 0.58 |
|  | Prefer not to answer | 0.11 (0.06, 0.20) | <0.001 | 4.38 (0.09, 207.68) | 0.45 |
|  | Other | 1.04 (0.12, 8.78) | 0.97 | 0.44 (0.04, 4.63) | 0.49 |
| Employment status | Full time/Part time | reference |  | reference |  |
|  | Maternity leave | 0.60 (0.39, 0.92) | 0.02 | 0.66 (0.41, 1.04) | 0.08 |
|  | Home duties | 0.75 (0.40, 1.38) | 0.35 | 0.98 (0.50, 1.92) | 0.96 |
|  | Unemployed | 0.41 (0.05, 3.35) | 0.41 | 1.15 (0.12, 12.27) | 0.91 |
|  | Student | 0.72 (0.17, 3.12) | 0.66 | 1.45 (0.18, 11.71) | 0.73 |
|  | Prefer not to answer | 0.04 (0.03, 0.07) | <0.001 | 1.48 (0.03, 70.35) | 0.84 |
| Subgroups | Future parent (no young children) | reference |  | reference |  |
|  | Current parent (young children) | 2.34 (1.25, 4.40) | 0.01 | 12.26 (2.82, 53.28) | <0.001 |
| **Personal experience with RSV** | Yes | 8.02 (3.99, 16.12) | <0.001 | 6.66 (3.26, 13.64) | <0.001 |
|  | No | *reference* |  | *reference* |  |
|  | Unsure | 1.28 (0.76, 2.15) | 0.36 | 1.15 (0.66, 1.99) | 0.63 |

\*\*All the variables depicted in Table 1 were entered into the model, however, were retained only if $p \leq 0.2$ . Other variables that were entered into the model but not shown in Table 1, were subgroups (pregnancy only, young child), and whether participants who currently have young children had previous experience with their child contracting an RSV infection. Education level 'other' refers to participants who may be involved in Vocational Education and Training (VET), foundation courses, English courses or Business and Management Education etc.
OR = odds ratio
a OR = adjusted odds ratio
CI = confidence interval
NA = not able to be estimated

Prior to being provided information on RSV, all pregnant/planning parents (including those with and without prior children) had a high level of acceptance (79.7%, n=370) towards receiving a maternal RSV vaccine. Similarly, the acceptance of a maternal vaccine was high in the future parent group (including only those without prior children; 86.9%, n=53). Overall, future (93.4%) and current parents (81.4%) had a high level of acceptance towards an infant RSV immunisation prior to information being provided.

There were no participants in the future parent subgroup and only 20 (1.0%) participants in the current parent subgroup that were not accepting of infant immunisation. Future parents who were accepting of both immunisation strategies (N=57) were asked on their immunisation strategy preference; majority accepted the maternal vaccine in combination or isolation of an infant immunisation as opposed to only an infant immunisation (both: 49.1%, maternal vaccine: 38.6%, infant immunisation: 3.5%).

Prior to completing the survey, few ‘future’ parents were unaware of RSV (10/61), but 90% (9/10) of these were willing to accept a maternal vaccine prior to learning about RSV, which increased to 100% after receiving information. Similarly, 97 of 1,931 ‘current’ parents were unaware of RSV prior to participation, however, 78.4% (76/97) were accepting of infant immunisation prior to learning about RSV, which increased to 85.6% (83/97) after receiving information.

The amount current and future parents were willing to pay for immunisations did not significantly differ before or after being provided with information on RSV. However, the median price of RSV childhood immunisation increased from A$69.00 to A$75.00 in future parents following information being provided (Figure 2).

**Figure 2.**
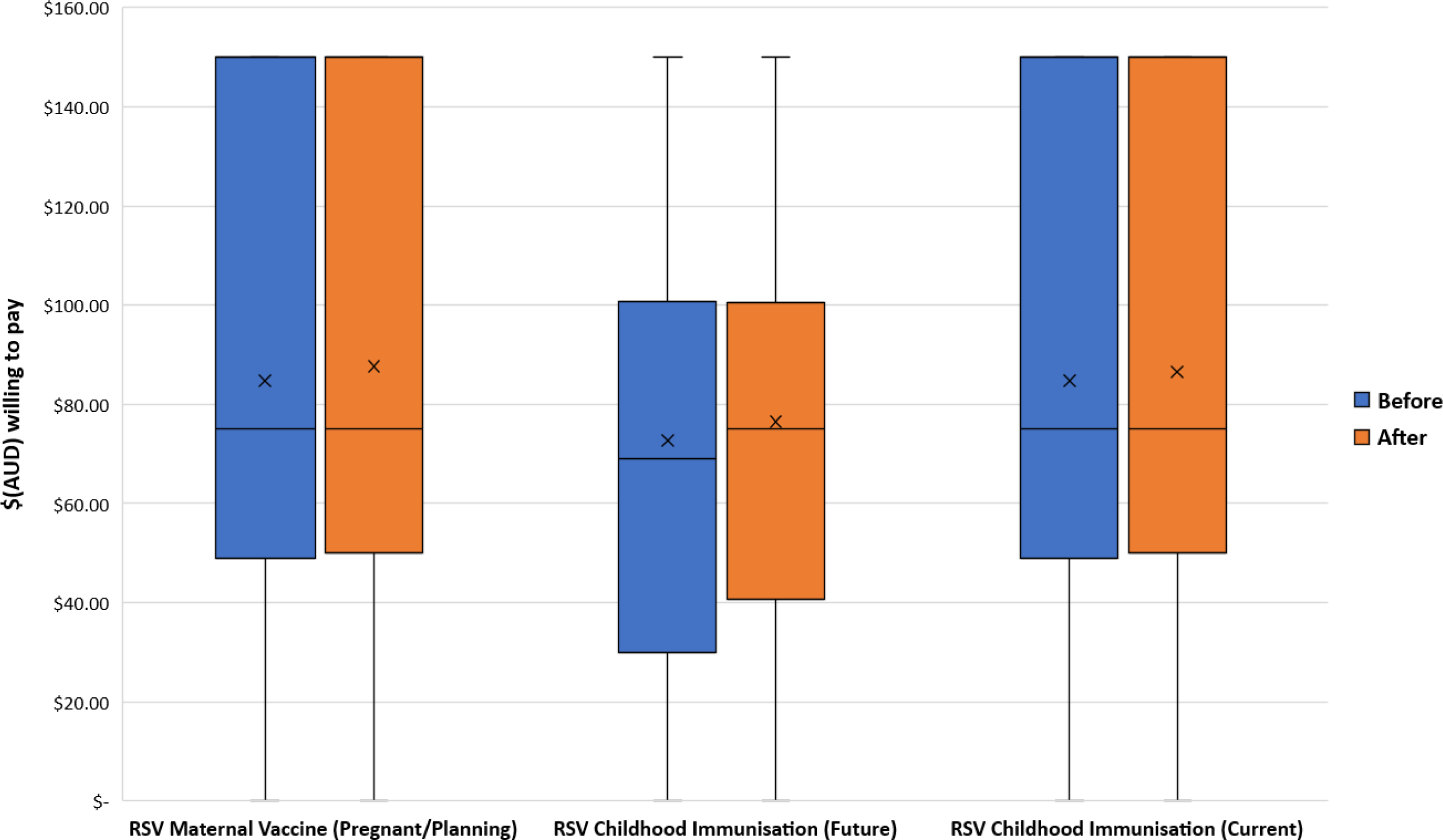
Amount parents were willing to pay for new RSV prevention strategies. Pregnant/planning parents were questioned on RSV maternal vaccines, and all parents (future and current) were questioned on RSV childhood immunisation. Participants were able to select any amount ranging from $0-$150. Note: excludes participants who did not respond.

Participants in the pregnant/planning subgroup identified their top 3 sources for infectious disease and vaccine information as their midwife (32.1%), google search (26.3%) and obstetrician (14.0%). Of those who selected google search, nearly half (46.0%) further indicated Government websites, 28.3% hospital websites, 12.4% science articles and 9.7% special health websites. Participants in the current parent subgroup identified their top 3 sources for information as google search (34.6%), their nurse (21.0%) and midwife (13.6%). Similar, most common google searches were for special health websites (44.4%), hospital websites (40.7%), Government websites (7.8%) or science articles (6.1%).

From 1,538 participants responding to the open-ended question regarding information needs for immunisation decision-making, three key themes were identified: defining RSV, prevention product information, and research and evidence (having peer-reviewed scientific evidence). Participants highlighted they need further information on RSV before accepting immunisations, including understanding the virus, symptoms, complications, severity, likelihood of infection and age groups most at risk. Respondents indicated that they’d also require information on the specific RSV prevention product, including benefits and risks, effectiveness, number and frequency of doses, management of side effects, and ideal age/s of administration. Some respondents indicated the need for approval from relevant government bodies and medical authorities, results and safety data from clinical trials and information from peer-reviewed sources.

## Discussion

We present here contemporary data on the parental awareness of RSV and acceptability of RSV immunisation strategies, likely to be available in Australia in the next 1-2 years. Current and future parents had a high level of awareness of most common childhood respiratory illnesses, including RSV. Notably, current parents were more aware of RSV compared to future parents. Though most participants had heard of RSV, there was a lack of awareness on associated conditions, especially among future parents. Indeed, the strongest independent factor of RSV awareness was being a current parent of a child aged ≤5 years.

Contrary to our findings, an earlier study in 2019 in Australia found majority of participants (83.0%) had never heard of RSV.^17^ However, our results are similar to that observed in a recent global study; majority of participants were only aware of the name of the virus and experienced parents had a higher level of awareness compared to new parents.^18^ The global study shared other similar observations; participants were more concerned about pneumonia than RSV perse and were more concerned about RSV than they were about bronchiolitis.^18^ There are several factors that are likely to contribute to the high level of awareness of RSV observed in our study. Firstly, our survey was administered at a time of peak RSV activity in 2022, especially in New South Wales (1000 cases recorded in the third quarter of 2022)^22^ and Victoria (11,291 cases in the same period).^22^ Secondly, there was increased media coverage on RSV in Australia due to the heightened number of cases^23^, and thirdly, our survey demographic was a highly educated population. Regardless of the high proportion of participants who were aware of RSV, the knowledge of key associated conditions including pneumonia and bronchiolitis was significantly low. This suggests the need for more education amongst Australian parents, particularly first-time parents, on disease severity of RSV in children.

In our study, parents had a high level of acceptance towards both RSV immunisation strategies prior to receiving information. Additionally, parents who had never heard of RSV became more accepting of immunisation once they received information. This demonstrates the importance of adequate knowledge of RSV as a key contributing factor towards acceptability of RSV immunisation. Furthermore, the median price future parents were willing to pay for RSV childhood immunisation increased by A$6 following receiving information. This conjecture was supported by both the Melbourne^17^ and global^18^ study. Both current and future parents referred to reputable and reliable sources for information regarding infectious diseases and vaccines. Furthermore, the qualitative findings in the study indicate that parents require information on RSV, as well as on associated prevention products and evidence supporting their use, to help in decision-making for RSV immunisations. Thus, when RSV immunisation strategies are implemented in Australia, it is vital that parents are equipped with information on the product and the disease. Such materials should particularly be targeted towards future parents; there is evidently a critical time and opportunity to inform new parents about RSV.

A major strength of our study was the large sample size we were able to attain through online recruitment. Facebook was the social media platform that reached the highest level of engagement with 159,624 impressions, 84 comments, 37 shares and 128 reactions. The use of a broad message summarising the purpose of the study, an engaging image (e.g., infant or pregnant mother receiving an immunisation), and the survey accessible online, were important factors to the successful participant recruitment. A further strength of our study was the co-designing of questions with community members including question wording and how to make the survey advertising appeal to the target audience. The result of community involvement in our research produced a simple, inclusive and easy to understand survey.

Despite these strengths, our study was not without limitations. The narrow demographic range of participants significantly reduced the generalisability of the results to the overall Australian population.^21^ People who are highly educated, spoke English, and from a middle to upper socioeconomic class were over-represented in the data. For this reason, there may have been an overestimation of the level of RSV awareness and acceptance of future immunisations. Another limitation of the study was that the online promotion of the survey was targeted to those engaged online with organisations involved in awareness of childhood respiratory conditions. This may have increased selection bias in the study as there is an assumption that participants who are engaged online with such organisations would have a heightened awareness about childhood respiratory conditions.

Further work investigating parental awareness and attitudes towards RSV prevention in Australia is needed, particularly focusing on groups from lower socio-economic backgrounds and targeting those who are at higher risk of severe outcomes associated with RSV. This could include groups with low levels of education, high unemployment rates, those from rural or remote Australia, Aboriginal and/or Torres Strait Islander families, culturally and linguistically diverse families, premature infants, and children with comorbidities. We are planning on conducting culturally appropriate research targeted towards and working alongside Aboriginal and/or Torres Strait Islander families.

## Conclusion

With Australia expected to follow Europe and the UK with licensure of mAb and expected licensure of a RSV maternal vaccine, more awareness of RSV among the community is required. Education and awareness campaigns would be most beneficial prior to childbirth, particularly among first time parents.

## Data Availability

Raw data produced in the present study is not available. However, aggregated data is available on request.

## Acknowledgements

The study investigators would like to acknowledge the community members and participants who contributed to the study, as well as Dr Monica Rikard-Bell who provided us with the survey questionnaire that was used in a COVID-19 vaccine acceptance study^19^ that we modified for our study. This study was funded through a Wesfarmers Centre of Vaccine and Infectious Diseases Seed Grant at the Telethon Kids Institute. HCM is funded by a Stan Perron Charitable Foundation Fellowship and a Future Health Research and Innovation Fund through the WA Near-miss Awards program.

## Conflicts of Interest

Associate Professor Richmond and Associate Professor Moore is in receipt of research funds from Merck Sharp & Dohme (Australia) Pty Ltd and Sanofi-Aventis Australia Pty Ltd (unrelated to the work presented in this paper). Associate Professor Richmond and Associate Professor Moore has also received institutional honoraria for participating in advisory committees (Pfizer, Merck Sharp & Dohme, EvoHealth) also unrelated to the work presented in this paper.

## Author contributions

Holland C had primary responsibility for survey development, statistical analysis (quantitative) and writing the manuscript.

Baker M had responsibility of project management, statistical analysis (qualitative) and editing/reviewing the manuscript.

Bates A and Hughes C had investigator roles and responsibility of providing feedback on the survey questionnaire and design and editing/reviewing the manuscript.

Dr Carlson and A.Prof Moore HC had primary responsibility of project conceptualisation, protocol development, methodology development, supervision and editing/reviewing the manuscript.

A.Prof Richmond supervised the overall project.

---

**Table 1.** Awareness of conditions associated with RSV. Analysis includes only those participants who advised that they had heard of the virus before prior to completing the survey (N=1779).

| Condition | Response | Future Subgroup |  | Current Subgroup |  | p-value |
| --- | --- | --- | --- | --- | --- | --- |
|  |  | N=48 |  | N=1731 |  |  |
|  |  | n | (%) | n | (%) |  |
| Influenza (the flu) | Yes | 13 | 27.1% | 434 | 25.1% | 0.52 |
|  | No | 16 | 33.3% | 641 | 37.0% |  |
|  | Unsure | 12 | 25.0% | 309 | 17.9% |  |
|  | Did not respond | 7 | 14.6% | 347 | 20.0% |  |
| Whooping cough | Yes | 4 | 8.3% | 143 | 8.3% | 0.47 |
|  | No | 26 | 54.2% | 825 | 47.7% |  |
|  | Unsure | 11 | 22.9% | 340 | 19.6% |  |
|  | Did not respond | 7 | 14.6% | 423 | 24.4% |  |
| Chest infection | Yes | 28 | 58.3% | 1188 | 68.6% | 0.081 |
|  | No | 6 | 12.5% | 139 | 8.0% |  |
|  | Unsure | 12 | 25.0% | 248 | 14.3% |  |
|  | Did not respond | 2 | 4.2% | 156 | 9.0% |  |
| Pneumonia** | Yes | 24 | 50.0% | 1112 | 64.2% | 0.009 |
|  | No | 7 | 14.6% | 155 | 9.0% |  |
|  | Unsure | 15 | 31.3% | 278 | 16.1% |  |
|  | Did not respond | 2 | 4.2% | 186 | 10.7% |  |
| Wheezing** | Yes | 27 | 56.3% | 1293 | 74.7% | 0.003 |
|  | No | 7 | 14.6% | 90 | 5.2% |  |
|  | Unsure | 11 | 22.9% | 214 | 12.4% |  |
|  | Did not respond | 3 | 6.3% | 134 | 7.7% |  |
| Bronchiolitis** | Yes | 25 | 52.1% | 1242 | 71.8% | 0.014 |
|  | No | 7 | 14.6% | 114 | 6.6% |  |
|  | Unsure | 13 | 27.1% | 273 | 15.8% |  |
|  | Did not respond | 3 | 6.3% | 102 | 5.9% |  |
| COVID-19 | Yes | 2 | 4.2% | 125 | 7.2% | 0.095 |
|  | No | 24 | 50.0% | 882 | 51.0% |  |
|  | Unsure | 14 | 29.2% | 287 | 16.6% |  |
|  | Did not respond | 8 | 16.7% | 437 | 25.2% |  |
| **Common cold** | Yes | 18 | 37.5% | 733 | 42.3% | 0.14 |
|  | No | 12 | 25.0% | 471 | 27.2% |  |
|  | Unsure | 13 | 27.1% | 261 | 15.1% |  |
|  | Did not respond | 5 | 10.4% | 266 | 15.4% |  |
| **Asthma\*\*** | Yes | 15 | 31.3% | 530 | 30.6% | 0.29 |
|  | No | 15 | 31.3% | 530 | 30.6% |  |
|  | Unsure | 13 | 27.1% | 343 | 19.8% |  |
|  | Did not respond | 5 | 10.4% | 359 | 20.7% |  |
| **Ear infections** | Yes | 8 | 16.7% | 366 | 21.1% | 0.16 |
|  | No | 18 | 37.5% | 621 | 35.9% |  |
|  | Unsure | 16 | 33.3% | 373 | 21.5% |  |
|  | Did not respond | 6 | 12.5% | 371 | 21.4% |  |
| **Gastroenteritis (gastro)** | Yes | 5 | 10.4% | 93 | 5.4% | 0.13 |
|  | No | 23 | 47.9% | 864 | 49.9% |  |
|  | Unsure | 13 | 27.1% | 337 | 19.5% |  |
|  | Did not respond | 7 | 14.6% | 437 | 25.2% |  |
| **Meningitis** | Yes | 6 | 12.5% | 110 | 6.4% | 0.13 |
|  | No | 21 | 43.8% | 796 | 46.0% |  |
|  | Unsure | 14 | 29.2% | 392 | 22.6% |  |
|  | Did not respond | 7 | 14.6% | 433 | 25.0% |  |

\*\* refers to conditions that are associated with RSV.

## Notes

### Author Declarations

Ethics approval was obtained by the University of Western Australia Human Research Ethics Committee (ET000182).

